# Clinical characteristics and prognosis in patients with acute myocarditis or unexplained acute chest pain: a nationwide longitudinal cohort study

**DOI:** 10.1101/2024.06.27.24309618

**Authors:** Marie Björkenstam, Emanuele Bobbio, Christian L. Polte, Clara Hjalmarsson, Niklas Bergh, Elmir Omerovic, Entela Bollano

**Author notes:** Correspondence to: Marie Björkenstam, MD, Department of cardiology, Sahlgrenska University Hospital, 41345 Gothenburg, Sweden.

## Abstract

**Background:** Acute myocarditis is a disease with variable prognosis, ranging from complete recovery to end-stage heart failure (HF) and death. The aim of this study was to examine the risk of mortality and HF in patients with suspected acute myocarditis (AM) in comparison to patients hospitalized for unexplained acute chest pain (ACP).

**Methods:** We used the SWEDEHEART-registry to identify patients >16 years admitted to hospital between 1 January 1998 and 31 December 2018 with either AM or ACP. Risks of all-cause mortality and development of HF were calculated and compared by use of adjusted Cox regression analyses.

**Results:** A total of 3,792 patients with AM and 109,934 patients with ACP were included. Median follow-up time was 7.8 years (Q1, Q3; 3.4, 12.3). AM patients were younger compared to ACP patients, median age 37 years (Q1, Q3; 26, 52) vs 59 years (Q1, Q3; 49, 69), and more likely to be men (79.9% vs 51.4%, p <0.001). Comorbidity burden was lower within the AM cohort. Chest pain was the most common presenting symptom in both groups. Mortality rate at 30 days (OR 3.75, 95% CI 1.9-7.3, p<0,001) as well as long term (OR 2.0, 95% CI 1.69-2.39, p <0.001) were significantly higher among AM patients and AM patients were more likely to develop HF during follow-up (OR 2.3, 95% CI 1.81-2.93, p<0,001).

**Conclusions:** AM patients had a worse short -and long-term outcome compared with ACP. The risk for development of HF was higher for AM patients even after the first year.

## Introduction

Acute myocarditis (AM) is an inflammation of the heart muscle due to infectious, toxic, or autoimmune processes^1^. The clinical presentation of the disease ranges from non-life-threatening symptoms such as chest pain, fever, myalgia, palpitations, or exertional dyspnoea, to fulminant haemodynamic collapse and sudden death^2 3^. Long-term outcomes can vary depending on the severity of the initial inflammation and the extent of damage to the heart muscle, ranging from complete recovery to development of advanced heart failure (HF), need for heart transplantation or death^2^.

Acute myocarditis often mimics acute coronary syndrome, manifesting with sudden onset of chest pain and electrocardiogram abnormalities^4^. Distinguishing between the two on presentation is often challenging and frequently requires invasive assessment of the coronary arteries. Previous studies on the subgroup of patients with symptoms suggestive of myocardial infarction are limited to case reports and case series on young males^5^. Culprit free coronary angiography is found in 5-13% of patients with suspected acute coronary syndrome^6^, with a majority of them eventually receiving a diagnosis of myocarditis^7^. Another group of patients presenting with acute chest pain and culprit-free coronary angiography are classified as unexplained acute chest pain (ACP). ACP is usually described as sudden and severe chest discomfort that lacks an obvious cause, the diagnosis is mostly applied once alternative explanations (such as acute coronary syndrome) have been ruled out. Previous studies have shown that these patients are at increased risk of cardiovascular events^8-10^. Both AM and ACP have better prognosis than acute myocardial infarction.^11^ However, there is limited data comparing the two groups.

Previous research has shown that men are more susceptible to myocarditis than women, especially at a younger age ^12^ . There are limited comprehensive epidemiological studies investigating the presenting characteristics of acute myocarditis patients. We used data from the national registry SWEDEHEART (Swedish Web System for Enhancement of Evidence-Based Care in Heart Disease Evaluated According to Recommended Therapies) which encompasses all patients admitted to hospitals with coronary care units (CCU) in Sweden, to characterize patients with AM in comparison with ACP and to investigate the prognosis in each group.

## Methods

### Database and Patient selection

The SWEDEHEART registry continuously registers all patients admitted to hospitals with CCU; RIKS-HIA (Register of Information and Knowledge about Swedish Heart Intensive Care Admissions) is one component of the SWEDEHEART registry. The full protocol has been published previously^13^ and detailed information and the complete protocol are available online at http://www.ucr.uu.se/swedeheart/. On admission, patients receive written information about SWEDEHEART and other quality-of-care registries; patients are permitted to deny participation in the registry, although few of them exercise this right. According to Swedish law, written consent is not required because quality control is an inherent element of hospital and other care. Research based on the registry is approved by an institutional ethics committee and all personal identifiers are removed from the SWEDEHEART data file when used for research purposes.^14^ RIKS-HIA started in 1995 with 19 participating hospitals across the country; by 2008, all Swedish hospitals with a CCU were participating in the registry.

Detailed information on approximately 100 variables is reported in case records during the hospitalisation period and is filled in by nurses^13^. In 2009 SWEDEHEART was founded by the merging of RIKS-HIA and three other registers^13^. Source data have continuously been validated by an external monitor via comparison of the information in the registry with hospital patient records. A 94% agreement was observed between the registered information and the source data in patients’ records, comprising 161 280 data points from 38 hospitals ^15^.

Diagnoses of diabetes, hypertension, hyperlipidemia, previous myocardial infarction, or previous stroke are made according to the International Classification of Diseases codes ^16^. Long-term survival data were obtained by merging the SWEDEHEART database with the Swedish Cause of Death Register, based on the patient’s unique 10-digit personal identification number^13^.

### Validation of the diagnosis

Studies that have assessed the accuracy of acute myocarditis diagnosis based on hospital record reviews have reported a correctness rate of over 80% in previous publications^17^.

### Definition and outcomes

The present study included all consecutive patients aged >16 years admitted between January1^st^, 1998 and December 31^st^, 2018, and reported to SWEDEHEART. Diagnoses were coded at the treating physician’s discretion according to the International Classification of diseases version 10 (ICD-10). AM was defined as a discharge diagnosis with one of the following main ICD codes: I40.0, I40.1, I40.8, I40.9 I41.0, I41.1, I41.2, I41.8 or I01.2. ACP was defined as discharge diagnosis R07.4.

The primary outcome was all-cause mortality at 30 days and long-term (median follow-up time 7.8 years). The risk of myocardial infarction, development of HF, significant bleeding and stroke was also studied. Patients were followed until death, emigration, or end of follow-up (December 2022).

### Statistics

Continuous variables are reported as mean ±standard deviation or as median and quartile (Q1-Q3), according to normal or non-normal distribution. Students t-test or Mann-Whitney U-test was used to compare continuous variables, as deemed appropriate. Kaplan-Meier curves were compared with the use of log-rank statistics. However, the underlying assumption of proportional hazards in the Cox model through follow-up was not met (treatment-time interaction, p <0.001). Comparisons between groups were therefore performed by multivariable adjusted logistic regression with follow-up time included as log-transformed offset variable, with the use of an estimated standard error for the difference. Differences with values of p <0.05 were considered statistically significant. Software packages used were Stata 18 and R (version 4.3.1).

### Ethics

The study was approved by the Swedish Ethical Review Authority (Dnr 2021-04026) and is in compliance with the Declaration of Helsinki.

## Results

The study population comprised 3,792 patients with AM and 109,934 patients with unexplained ACP. The baseline characteristics are shown in **table 1**. Patients with AM were younger (median age 37 (Q1, Q3; 26, 62) vs 59 years (Q1, Q3; 49, 69), more often male, and had a lower burden of common comorbidities compared to patients with ACP. The prevalence of diabetes was 4.7% and 8.7% in the AM and ACP groups, respectively (p-value < 0.001) , while hypertension was found in 3.7% and 12.7% of patients of AM and ACP, respectively (p-value < 0.001)

**Table 1:** Patients characteristics at admission to the cardiac care unit.

|  | **Acute myocarditis** | **Unexplained chest pain** | **p-value** |
| --- | --- | --- | --- |
| **N** | 3792 | 109934 |  |
| **Age (median (IQR))** | 37.00 (26.00, 52.00) | 59.00 (49.00, 69.00) | <0.001 |
| **Male, n (%)** | 3028 (79.9%) | 56533 (51.4%) | <0.001 |
| **Female, n (%)** | 764 (20.1%) | 53401 (48.6%) | <0.001 |
| **Smoking (%)** |  |  | <0.001 |
| **Never smoker** | 58.5% | 49.3% |  |
| **Past smoker** | 17.6% | 27.8% |  |
| **Smoker** | 20.2% | 19.5% |  |
| **Diabetes, n (%)** | 152 (4.0%) | 9554 (8.7%) | <0.001 |
| **Hypertension, n (%)** | 140 (3.7%) | 13964 (12.7%) | <0.001 |
| **Previous stroke, n (%)** | 49 (1.3%) | 4336 (3.9%) | <0.001 |
| **Previous HF, n (%)** | 46 (1.2%) | 2626 (2.4%) | <0.001 |
| **Previous cancer (%)** | 34 (0.9%) | 1300 (1.2%) | 0,11 |
| **Previous CABG (%)** | 10 (0.3%) | 1812 (1.6%) | <0,001 |
| **Previous PCI (%)** | 8 (0.2%) | 1429 (1.3%) | <0,001 |

HF, heart failure; CABG, coronary artery by-pass grafting; PCI, percutaneous coronary intervention

The most common presenting symptom for patients with AM was chest pain (82.9%), followed by dyspnea (3.1%). Chest pain was also the most common presenting symptom in the ACP group (93.4%). In our material, a very small proportion presented with cardiac shock or resuscitated cardiac arrest, but the rate of cardiac shock/resuscitated cardiac arrest was significantly higher in the AM group relative to the unexplained ACP group (0.6% and 0.2% respectively for cardiac shock and 0.7% vs <0.1% for resuscitated cardiac arrest, p-value <0.001). Compared with patients with unexplained ACP, AM patients displayed higher levels of cardiac troponin [10 (0.92, 127) ng/L vs 0.03 (0.01, 2) ng/L; p-value <0.001] as well as C-reactive protein (CRP) [29 (6, 75) mg/L vs 5 (2.2, 8) mg/L; p-value <0.001] (**table 2**). ECG-changes in the form of ST-segment elevation were more common in patients with AM (43.5% vs 4.7%, p-value <0.001, see **table 3**).

**Table 2:** Presenting symptoms, clinical and laboratory findings at admission.

|  | **AM** | **ACP** | **p-value** |
| --- | --- | --- | --- |
| **N** | 3792 | 109934 |  |
| **SPB mmHg (median, IQR)** | 130 (120, 145) | 148 (130, 165) | <0,001 |
| **DBP mmHg (median, IQR)** | 80 (70, 90) | 84 (75, 93) | <0,001 |
| **Pulmonary rales (%)** |  |  | <0,001 |
| **No** | 95.6% | 97.4% |  |
| **Yes** | 3.1% | 1.9% |  |
| **Presenting symptoms (%)** |  |  | <0,001 |
| **Chest pain** | 82.9% | 93.4% |  |
| **Dyspnoea** | 3.1% | 1.1% |  |
| **Cardiac shock** | 0.6% | 0.2% |  |
| **Other symptoms** | 9.1% | 4.1% |  |
| **Resuscitated cardiac Arrest (%)** | 0.7% | <0.1% | <0,001 |
| **VT/VF** | 0.4% | <1% |  |
| **Laboratory findings** |  |  |  |
| **Troponins (median, IQR)** | 10.00 (0.92, 127.00) | 0.03 (0.01, 2.00) | <0.001 |
| **CRP (median, IQR)** | 29.00 (6.00, 75.00) | 5.00 (2.20, 8.00) | <0.001 |
| **b-glucose (median, IQR)** | 6.00 (5.40, 6.90) | 5.80 (5.20, 6.70) | <0.001 |
| **Hemoglobine (median, IQR)** | 144 (135, 152) | 141 (132, 150) | <0.001 |
| **HbA1c (median, IQR)** | 36 (34, 39) | 38 (35, 43) | <0,001 |
| **Creatinine (median, IQR)** | 78 (68, 89) | 75 (65, 88) | <0,001 |
| **Cholesterol (total) (median, IQR)** | 4.30 (3.70, 5.10) | 5.20 (4.50, 6.00) | <0,001 |

AM, acute myocarditis; ACP, acute chest pain; SBP, systolic blood pressure; DBP, diastolic blood pressure; CRP, C-reactive protein; HbA1c, hemoglobine A1c

**Table 3.** Electrocardiographic findings at admission and at discharge.

|  | AM, n (%) | ACP, n (%) | P-value |
| --- | --- | --- | --- |
| **ECG RHYTHM admission** |  |  | <0.001 |
| Sinus | 3156 (95.1%) | 91980 (96.5%) |  |
| Atrial fibrillation/flutter | 78 (2.4%) | 1868 (2.0%) |  |
| Other | 53 (1.6%) | 754 (0.8%) |  |
| **QRS COMPLEX** |  |  | <0.001 |
| Normal | 2780 (84.5%) | 78159 (83.0%) |  |
| Pacemaker | 9 (0.3%) | 479 (0.5%) |  |
| Left bundle branch block | 50 (1.5%) | 2586 (2.7%) |  |
| Pathologic Q-wave | 77 (2.3%) | 3267 (3.5%) |  |
| Right bundle branch block | 53 (1.6%) | 2100 (2.2%) |  |
| Other | 254 (7.7%) | 6164 (6.5%) |  |
| **ST-SEGMENT CHANGES** |  |  | <0.001 |
| Normal | 1036 (31.5%) | 64564 (68.7%) |  |
| ST-elevation | 1434 (43.5%) | 4406 (4.7%) |  |
| ST-depression | 167 (5.1%) | 7245 (7.7%) |  |
| Pathologic T-wave | 317 (9.6%) | 7762 (8.3%) |  |
| Other | 289 (8.8%) | 8620 (9.2%) |  |
| **DISCHARGE ECG** |  |  | 0.002 |
| Sinus | 2979 (95.5%) | 86626 (96.6%) |  |
| Atrial fibrillation/flutter | 50 (1.6%) | 1205 (1.3%) |  |
| Other | 29 (0.9%) | 466 (0.5%) |  |

AM, acute myocarditis; ACP, acute chest pain

Registration of medication at discharge from hospital showed that patients with AM were less often treated with Angiotensin-Converting Enzyme inhibitors than patients with unexplained ACP (4% vs 10.5 %; p-value <0.001). The same was true for Angiotensin receptor blockers (3.3% vs 8.8%; p-value < 0.001) and diuretics (3.8% vs 12.4%; p-value < 0.001). For more details on medications see **table 4**.

### Short time outcome (during hospitalization and at 30 days)

Patients with AM presented higher mortality at 30 days compared with unexplained ACP (OR 3.75, 95% CI 1.9-7.3, p<0.001) when analysed by multivariable adjusted logistic regression. The mortality at 30 days is shown in **Figure 1**.

**Figure 1.**
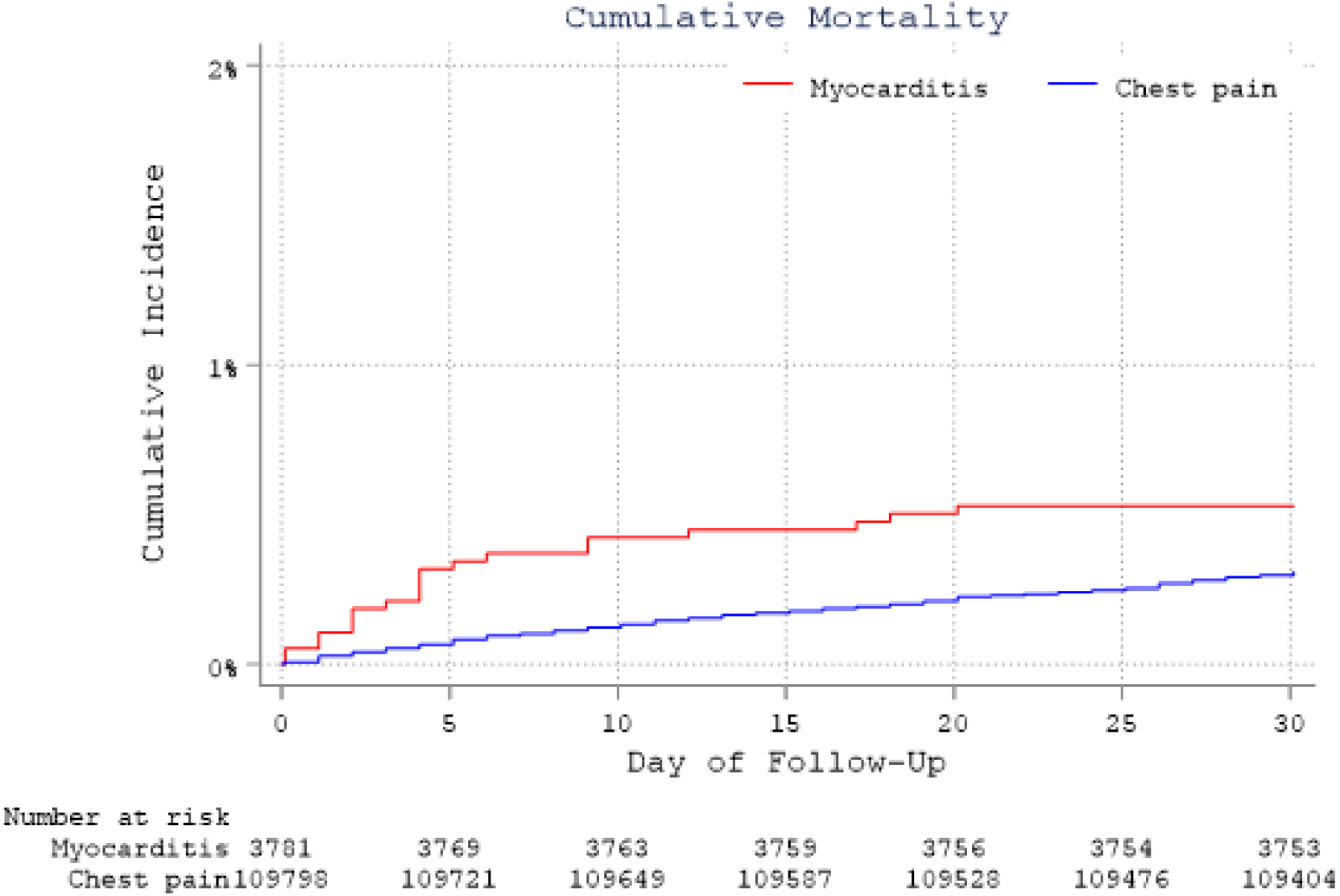
All cause mortality for patients with acute myocarditis and acute unexplained chest pain at 30 days

The risk of developing HF during hospitalization was increased for the AM cohort compared with patients with unexplained ACP (OR 5.4, 95% CI 4.44-6.49, p<0.001).

The number of patients admitted for AM undergoing coronary angiography increased with 3% per year during the study period (0.030, 95% CI 0.028-0.033, p<0.001, **Figure 2**) and during the last four years more than 60% had a coronary angiography performed to rule out acute coronary syndrome. In contrast, 10.7% of the unexplained ACP group underwent coronary angiography.

**Figure 2.**
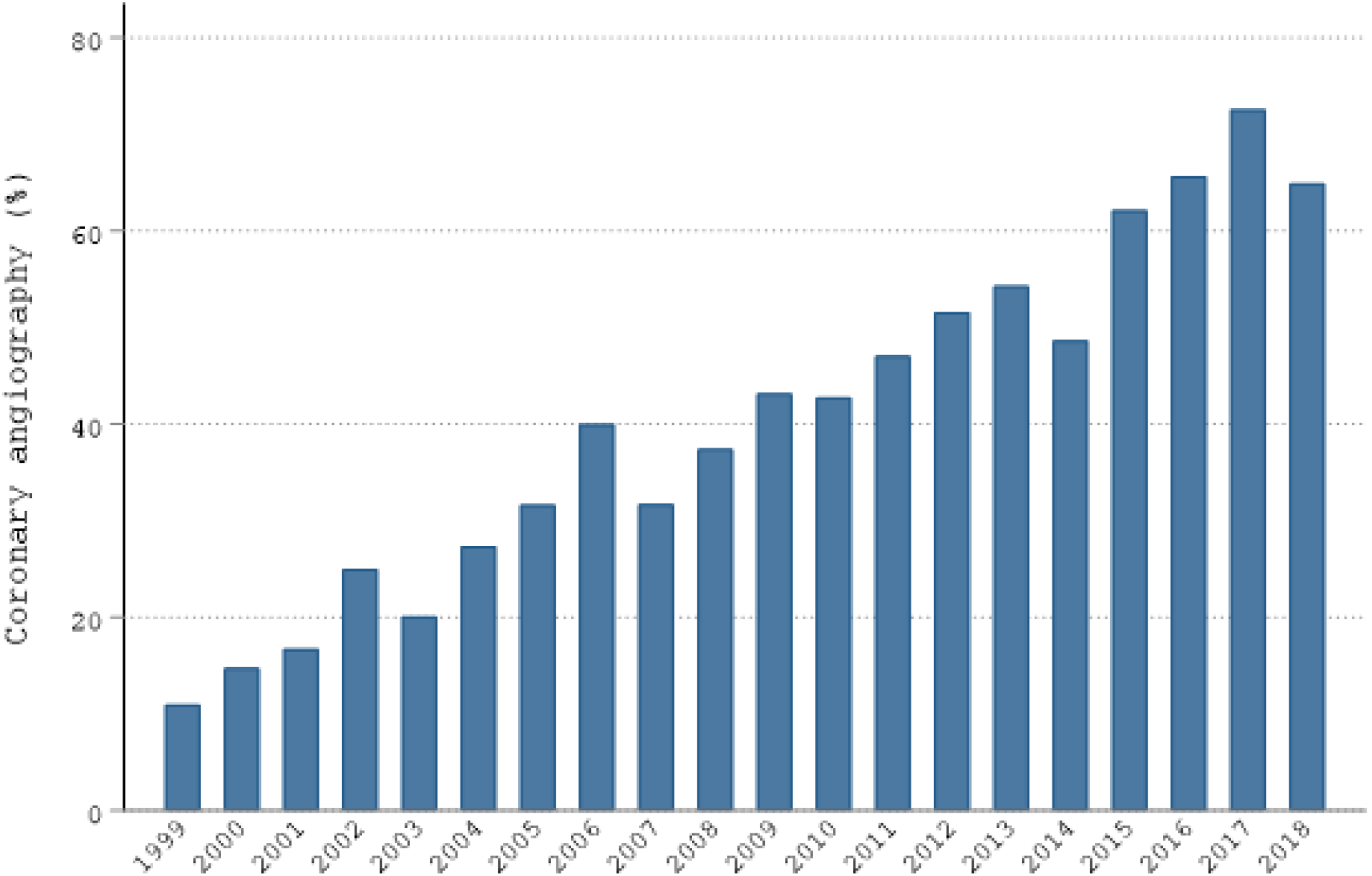
Percentage of patients hospitalized for suspected AM undergoing coronary angiography during 1999-2018

### Long term outcome

Under a median follow up of 7.8 years (Q1, Q3; 3.4, 12.3), patients with AM displayed worse survival (OR 2.0, 95% CI 1.69-2.39, p <0.001) compared with ACP, as well as higher risk for HF (OR 2.3, 95% CI 1.81-2.93, p<0.001) and myocardial infarction (OR 1.3, 95% CI 1.11-1.62, p< 0.001). No difference in the risk for stroke or bleeding was observed. The groups were compared using multivariable adjusted logistic regression.

## Discussion

This large, nationwide, longitudinal cohort-study showed that patients with AM, despite a younger age and less comorbidities, have worse short- and long-term survival compared with patients admitted due to unexplained ACP. Moreover, AM patients presented higher risk for developing HF both in-hospital and long-term after admission compared with unexplained ACP. These are important findings considering that AM is commonly regarded as a benign condition and shows the clinical importance of structured follow-up of these patients, both short term and long term.

Our study confirmed that patients with AM are usually young and predominantly male^2-4^. Recently, Ammirati et al. showed a very low mortality rate among patients with uncomplicated myocarditis^18^ which is in line with that AM is commonly considered a benign condition. While the absolute number of deaths in our study is small, it is noteworthy that the mortality risk was higher when compared with a group of patients who are both older and have more comorbidities. A possible explanation to this finding could be our inclusive analysis of all cases of AM rather than categorizing them into distinct groups such as uncomplicated and complicated cases, as previous studies have done^3 19 20^. The higher mortality is in line with previous studies reporting an increased risk of 1-year mortality in patients with myocarditis compared to healthy controls^17^.

The higher risk for HF short- and long-term in the AM cohort is interesting, particularly considering that patients with unexplained ACP tend to have a higher prevalence of co-morbidities in our cohort (e.g., hypertension, a common cause of HF^21^). This underscores the importance of monitoring AM patients, given the current variation in follow-up procedures among patients and medical centres. A study of patients discharged 1987-2006 with a diagnose of HF showed an increased hospitalization due to HF in young adults (aged 18-44 years) during the study period^22^. It is possible that part of that increase is caused by an AM leading to the development of HF. Previous studies have shown an association between myocarditis and the development of HF as well as dilated cardiomyopathy^23^. Our findings align with previous studies., which indicated a clinically significant long-term risk of HF and the need for pacing devices or ICD implantation following myocarditis compared to matched controls from the general population^24^. The same has also been reported in an earlier Swedish study^17^. The increased risk in the long-term for other cardiovascular events such as myocardial infarction in AM may be somehow related to inflammation, which is present in both myocarditis and unstable atherosclerotic plaques causing myocardial infarction^25-27^. Further research is needed to confirm and understand this relationship.

The increase in coronary angiographies in the AM group during the study period probably reflects changes in daily clinical practice and an increased availability. More patients undergoing coronary angiography may also be attributed to the introduction of high-sensitive troponins and thereby the possibility to detect lower levels of cardiac biomarkers which leads to more angiographies. Another possible contributing reason to the increase is the publication of the position paper on acute myocarditis from the European Society of Cardiology in 2013 which recommends the exclusion of coronary artery disease in patients with suspected myocarditis^2^.

### Strengths and Limitations

An important strength in this study lies in its nationwide design, encompassing all patients admitted to CCU in Sweden. Since it relies on a registry-based approach, there is minimal loss in follow-up data. There is also, to our knowledge, very limited previous research comparing these two groups of patients. Nonetheless, there are certain limitations to consider. Although the SWEDEHEART registry has undergone thorough validation, there always exists a risk of inaccurate data reporting within the registry. This is a retrospective analysis of registry data with the limitations inherent to such analyses. In clinical practice there is a challenge to distinguish between AM and pericarditis and patients diagnosed with pericarditis were not included in the analyzes. This may potentially result in missed cases of AM. Furthermore, diagnosing AM is notoriously complex, suggesting that this study may underestimate the actual incidence of hospitalizations for this condition. Furthermore, data collection of certain parameters, including left ventricular ejection fraction and cardiac magnetic resonance imaging findings, which would have been of interest, was not possible as it is not recorded in the SWEDEHEART registry.

## Conclusion

The findings of this study suggest that patients with AM have a greater risk of mortality at 30 days and long-term and a higher likelihood of developing HF compared with patients with unexplained ACP. Further investigation is required to create risk stratification strategies for AM patients.

## Data Availability

The data that support the findings of this study are available from the corresponding author upon reasonable request.

## Acknowledgments

No acknowledgments

